# Non-Invasive Detection of Biphasic Cardiac Troponin-I Release During and After Marathon Running Using Point-of-Care Saliva Analysis

**DOI:** 10.64898/2026.05.04.26352369

**Authors:** Aleksandr N. Ovchinnikov, Antonio Paoli

## Abstract

**Objectives:** This study aimed to compare exercise-induced changes in serum and salivary concentrations of cardiac troponin-I (cTnI) in athletes during and after a marathon.

**Methods:** Thirty-six male runners were recruited. Eighteen participants in group 1 completed a marathon (42.195 km), while eighteen participants in group 2 did not undergo this exercise. Blood and saliva samples were collected at twelve different time points and then analyzed for cTnI using an immunoassay.

**Results:** Biphasic cTnI release into the circulation was observed during and after the marathon. Moreover, a similar pattern of biphasic cTnI elevation was found in saliva. In group 1, salivary and serum concentrations of cTnI first peaked after 60 min of exercise (0.67±0.08 ng/mL and 0.76±0.07 ng/mL), decreased slightly towards the end of the marathon (0.40±0.06 ng/mL and 0.46±0.06 ng/mL), and then reached a second, higher peak 4 h post-exercise (0.72±0.09 ng/mL and 0.82±0.09 ng/mL), returning to baseline by 48 h after marathon completion (0.16±0.04 ng/mL and 0.18±0.04 ng/mL). In group 2, there were no time-dependent changes in cTnI concentrations in both saliva and serum. Deming regression and Passing–Bablok regression demonstrated that there was proportional agreement between salivary and serum levels of cTnI in both groups at all twelve time points. The Bland–Altman method revealed that there was a negative differential bias but no proportional bias in the data.

**Conclusions:** Documenting a similar, biphasic pattern of cTnI elevations in saliva and serum during and after the marathon provides a reliable non-invasive alternative without requiring a blood draw.

## Introduction

The presence of detectable cardiac troponin (cTn) in the circulation following strenuous exercise in the healthy population is a well-documented phenomenon [1–5]. Groundbreaking research by Middleton et al. demonstrated that marathon running causes a biphasic release of cardiac troponin-T (cTnT), with relatively minor elevations in the blood, indicating physiological and reversible rather than pathological and irreversible myocardial injury. Even though transient post-exercise cTn elevations are often observed in the circulation of exercising individuals, not all elevated states of cTn are made equal [3, 4, 6]. Sustained (>24 hours) post-exercise cTn elevation above the cut-off value (i.e., the 99th percentile of the upper reference limit) seems to be associated with increased risk of cardiac events [3, 5, 6]. Evidence exists that the risk of cardiac events, including sudden cardiac death, correlates with severity of myocardial inflammation and serum cTn levels [6, 7]. Therefore, repeated cTn quantifications are sometimes used for assessing cardiomyocyte damage after exercise-induced myocardial stress [3, 4].

When coupled with a suitable clinical scenario, cTn testing is the gold-standard for documenting acute myocardial injury [1–5, 8], but the current modality requires a blood draw [9]. Blood sampling, which is intrusive, logistically difficult, and inappropriate for dense, real-time monitoring, severely limits the availability and frequency of cTn testing, complicating immediate clinical decision-making [10]. Recent application of novel point-of-care (POC) technologies to alternative biofluids, such as saliva, opens new avenues for non-invasive, patient-friendly monitoring of cTn elevations [10–12]. Previous pivotal study revealed proportional agreement between salivary and serum concentrations of cardiac troponin-I (cTnI) before and after a 5-km time-trial using a POC immunoassay, validating saliva as a viable medium for identifying exercise-induced cTnI elevations [13]. However, this proof-of-concept research was restricted to one pre-exercise and three post-exercise time points for cTnI measurements in a relatively short-bout setting [13].

Here, we hypothesize that the biphasic cTn release during and after marathon running, previously reported only for cTnT in blood [14], may be non-invasively tracked in saliva. Although both cTnI and cTnT have been shown in numerous studies to be useful tools in detecting myocardial damage [1–5, 8, 9], quantifying cTnI in saliva may be more beneficial due to its molecular mass, which is significantly less than that of cTnT [13]. Consequently, it is logical to assume that the blood-salivary barrier is more easily crossed by cTnI. We also hypothesize that a POC cTnI assay utilizing saliva might provide essential resolution to map the kinetics of cTnI release during and after prolonged strenuous exercise, in particular the marathon. In order to test this, we used a validated POC device and a dense sampling protocol to measure salivary and serum concentrations of cTnI hourly during the marathon race and throughout recovery, comparing cTnI kinetics in saliva with those in blood serum.

## Materials and methods

### Participants and study design

A prospective, observational cohort study was conducted to compare exercise-induced changes in serum and salivary concentrations of cTnI during and after marathon running. Thirty-six well-trained male runners (age: 26.64±2.54 years; height: 174.64±3.05 cm; body mass: 64.97±3.01 kg; body mass index: 21.30±0.79 kg/m^2^) with at least six years of regional-level competition experience were recruited. All participants were healthy, asymptomatic, normotensive, non-smoking, and free of cardiovascular disease, as confirmed by the pre-participation screening examinations. Exclusion criteria for all volunteers included cardiovascular disease, chronic medication use, respiratory infection, orthopedic injury, fair or poor oral health, inability to provide a blood or saliva sample, and age less than 18 years.

The study was approved by the Local Ethics Committee (Nr. 2, 22.09.2025) and was conducted in accordance with the principles set forth in the Declaration of Helsinki [15]. Prior to enrollment, each volunteer provided written informed consent after being fully informed about the methodology and study design. After signing the informed consent form, all participants underwent medical screening in the university laboratories to determine eligibility. A qualified dental hygienist evaluated participants’ oral health in the dental office. Based on the degree of mucosal inflammation, the extent of decay and periodontal diseases, and the presence or absence of dental complaints, oral health was rated as poor, fair, or good by a single examiner using a modified version of the oral health scoring system [16]. Each participant was instructed to avoid ingesting alcohol and caffeine for at least 24 hours before saliva and blood collection and to eat breakfast 1 hour prior to the marathon race.

Each participant was a long-distance runner who followed a typical training regimen. Each training session lasted at least 2 hours, and there were at least 5 sessions per week. All participants refrained from exercise for 24 hours prior to the marathon run, which took place on a motorized treadmill in the morning. All participants were randomly assigned to two groups using computer-generated permuted blocks with a 1:1 ratio via sequentially numbered opaque envelopes in order to ascertain how marathon running might impact salivary and serum levels of cTnI. Of the 36 consecutive runners who were recruited, 18 participants in group 1 successfully completed a marathon (42.195 km), while 18 participants in group 2 (non-competitive controls) sat in chairs in the room close to the laboratory instead of competing in the marathon. A stopwatch was used to manually record the times of the runners in group 1, who were urged to exert maximum effort.

### Sample collection and cTnI measurement

Venous blood samples were drawn through a cannula and placed in vacutainers with anticoagulant (EDTA). Unstimulated saliva was collected into sterile tubes for 5 minutes using the spitting method. Before collecting saliva, all participants thoroughly rinsed their mouths and swallowed any remaining water. Saliva and blood samples were taken from participants in both groups at twelve different time points: at rest (pre-exercise), at 60-minute intervals during the marathon race, immediately after, and at 1, 2, 4, 6, 12, 24, and 48 hours after exercise. All samples were transferred on ice to a special core laboratory, where they were spun at 3000 rpm for 15 minutes, separated into aliquots, and stored at −80 °C until the cTnI measurement. Within three months of storage, serum and saliva were analyzed for cTnI using an immunoassay (Getein Biotech Inc., China), the detailed description of which was previously reported [13].

### Statistical analysis

The data are presented as mean ± standard deviation (SD). The median and interquartile range are used to express the data in Figure 1. The mean ± SD, augmented with the median and interquartile range, is used to express the data in Figure 2. A Shapiro–Wilk test was used to assess the assumption of normality. Since all the data were normally distributed, the Student’s t-test was used to analyze between-group differences, and the corresponding effect sizes were also calculated. An effect size of Cohen’s d = 0.97 would be required to ascertain between-group differences in cTnI levels in both saliva and serum, taking into account the sample size and a power of 80% with an α of 0.05. Since the Fligner–Killeen test results revealed that the variances were not homogeneous, within-group differences were examined using the Friedman test, with a Durbin–Conover pairwise post-hoc test in the event of significant differences. Pearson’s correlation coefficient was used to demonstrate a significant linear relationship between salivary and serum concentrations of cTnI. The ordinary least squares (OLS), Deming, and Passing–Bablok regression lines were shown in the scatter plots. Both a proportional bias (i.e., a difference that relies on the value of the mean of salivary and serum concentrations of cTnI) and a differential bias (i.e., a constant difference between salivary and serum concentrations of cTnI) were identified using the Bland–Altman method of differences. The Bland–Altman method’s assumptions were satisfied since the differences between salivary and serum concentrations of cTnI were normally distributed. A p-value less than 0.05 was deemed significant. If necessary, the p-value was adjusted using the Holm correction. RStudio (version 2022.07.2+576 for macOS, RStudio, PBC, Boston, MA; http://www.rstudio.com) was used for all statistical analyses.

**Figure 1:**
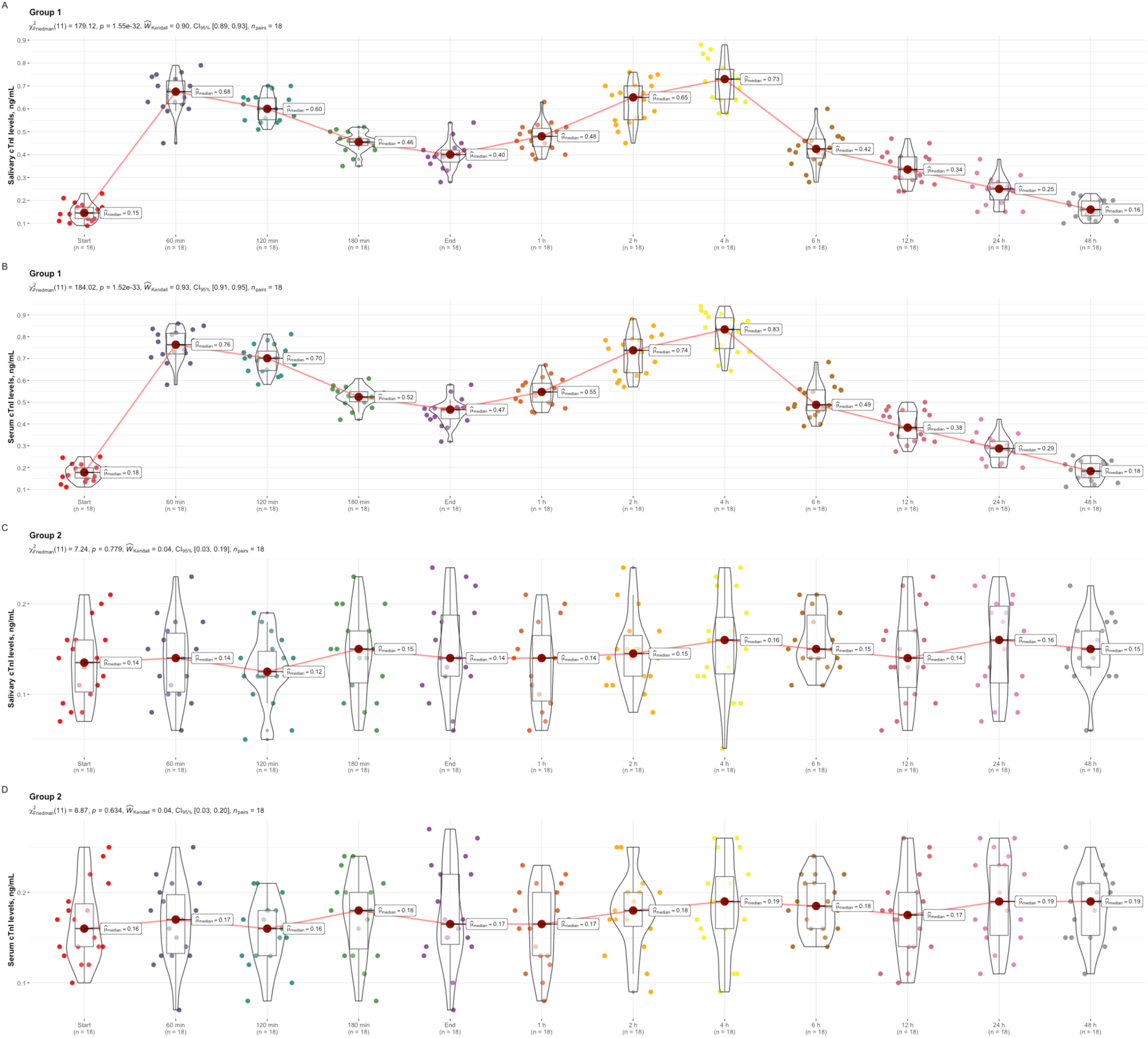
Salivary and serum levels of cTnI in participants of group 1. (A, B) and group 2 (C, D) during (min) and after (h) marathon running. Data are expressed as median and interquartile range, and are compared using the Friedman test, with a Durbin–Conover pairwise post-hoc test in the event of significant differences. cTnI, cardiac troponin-I.

**Figure 2:**
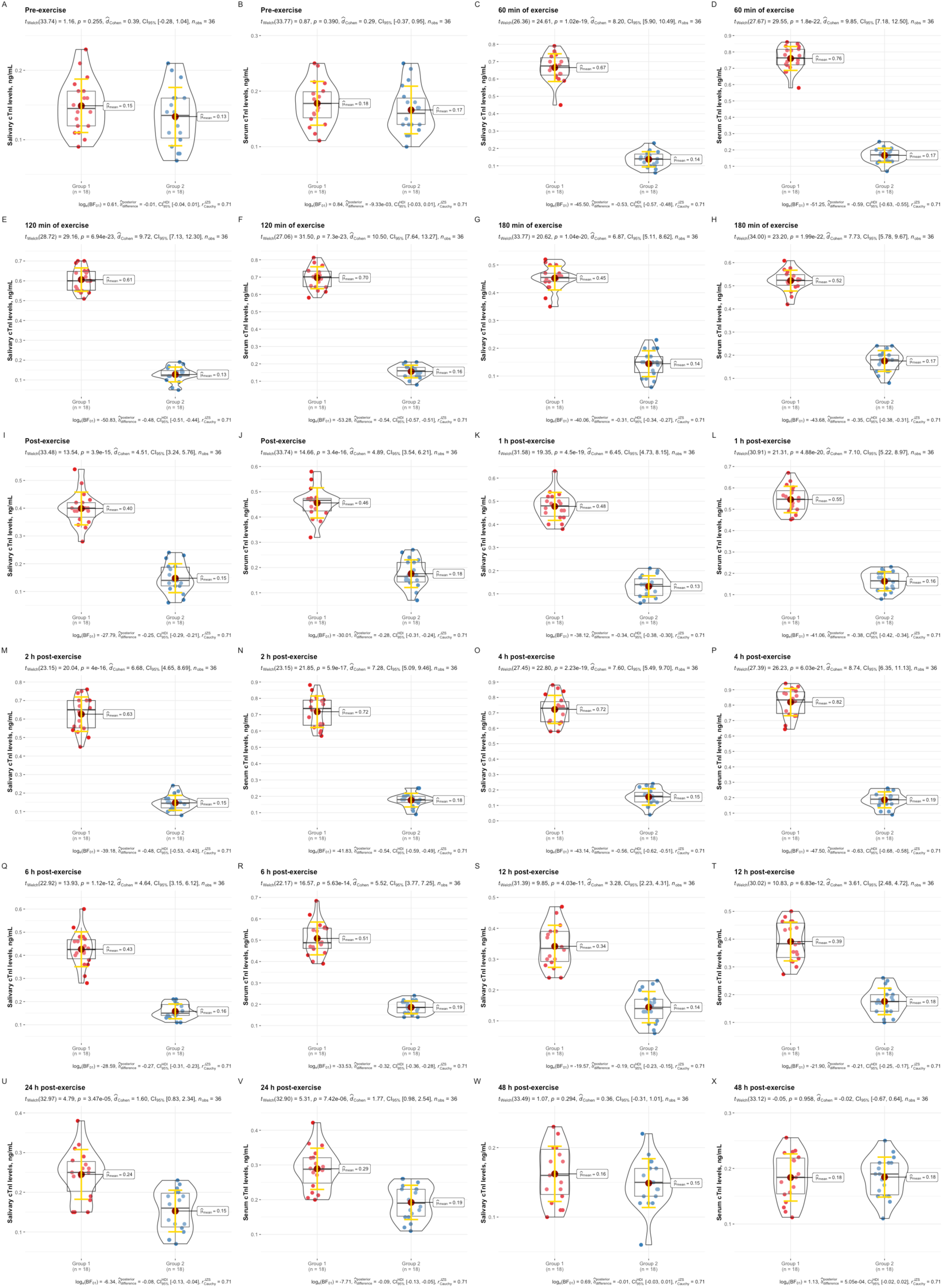
Salivary and serum levels of cTnI in participants of both groups at pre-exercise. (A, B), 60 min of exercise (C, D), 120 min of exercise (E, F), 180 min of exercise (G, H), immediately post-exercise (I, J), and at 1 h (K, L), 2 h (M, N), 4 h (O, P), 6 h (Q, R), 12 h (S, T), 24 h (U, V), and 48 h (W, X) post-exercise. Data are expressed as mean ± SD, with median and interquartile range shown, and were compared using Student’s t-test for independent samples. cTnI, cardiac troponin-I.

## Results

All participants tolerated the study well, and no adverse events were reported during and after marathon running in group 1. Times to complete the marathon (42.195 km) for the runners in group 1 were 215.21 ± 8.17 minutes.

Salivary and serum levels of cTnI were longitudinally evaluated across twelve time points during and after marathon running in both groups. Biphasic cTnI release into the circulation was observed during and after the marathon. Moreover, a similar pattern of biphasic cTnI elevation was found in saliva. In group 1, as shown in Figure 1, salivary and serum concentrations of cTnI first peaked after 60 minutes of exercise, decreased slightly towards the end of the marathon, and then reached a second, higher peak 4 hours post-exercise, returning to baseline by 48 hours after marathon completion.

In participants of group 2 who sat in chairs in the room close to the laboratory instead of competing in the marathon, there were no time-dependent changes in cTnI levels in both saliva and blood serum up to 48 hours after the marathon. Salivary and serum concentrations of cTnI did not differ between groups at pre-exercise and 48 hours post-exercise, but were significantly lower in group 2 compared to group 1 at all other time points (Figure 2).

A significant positive correlation was observed between cTnI levels in saliva and serum in both groups at all twelve time points (Figure 3).

**Figure 3:**
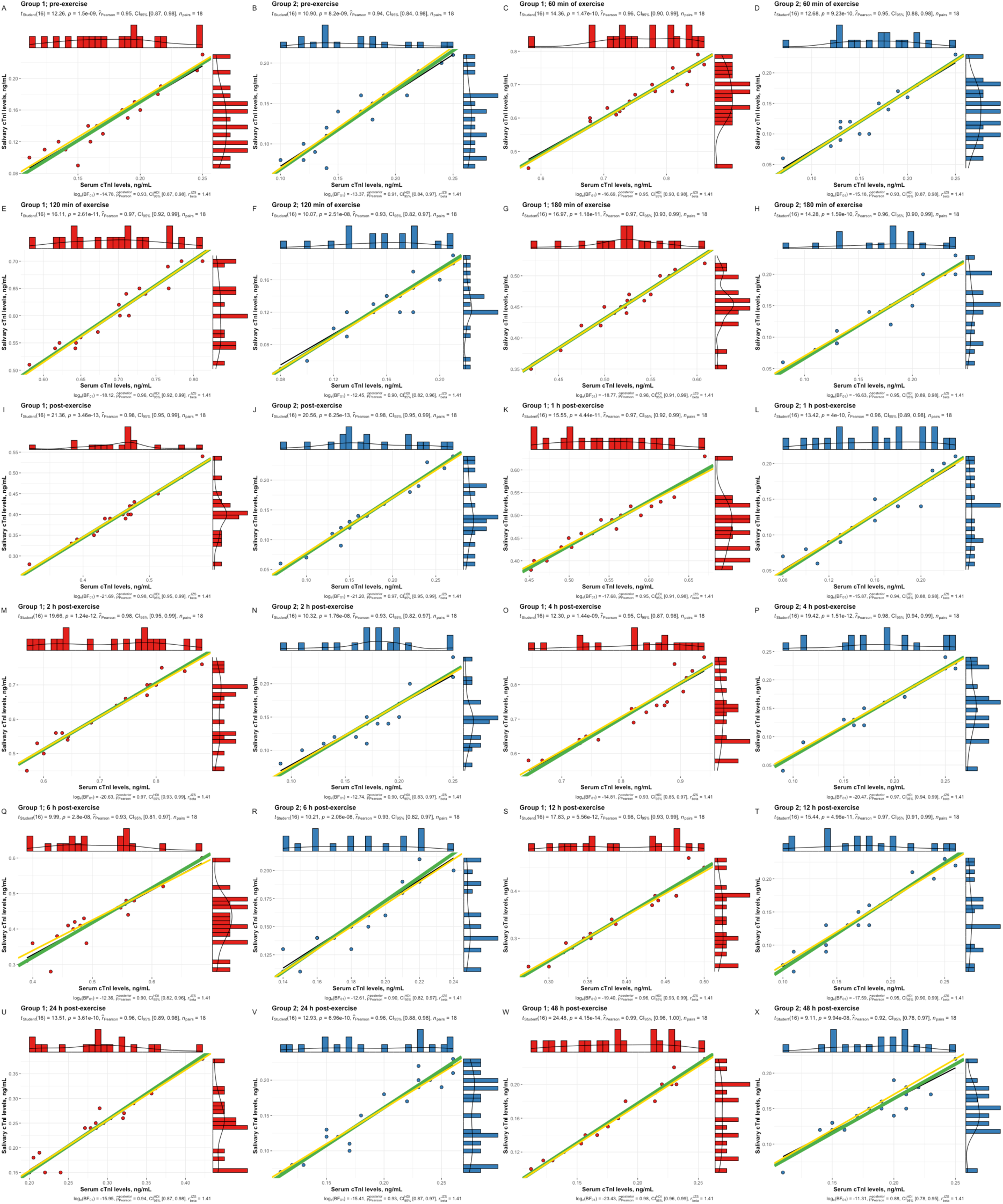
Correlation between salivary and serum levels of cTnI in participants of group 1. (A, C, E, G, I, K, M, O, Q, S, U, W) and group 2 (B, D, F, H, J, L, N, P, R, T, V, X) during and after marathon running. The solid black lines represent the ordinary least squares regression of salivary cTnI levels on serum cTnI levels. The solid green and yellow lines represent the Deming regression and Passing–Bablok regression, respectively. cTnI, cardiac troponin-I.

Deming regression and Passing–Bablok regression demonstrated that there was proportional agreement (i.e., the 95% confidence interval (CI) for the slope contained the value 1) between salivary and serum levels of cTnI in both groups at all twelve time points.

The Bland–Altman method of differences indicated that there was a negative differential bias in the data because the line of equality was higher than the 95% CI of the mean difference between salivary and serum levels of cTnI in participants of both groups at all twelve time points (Figure 4).

**Figure 4:**
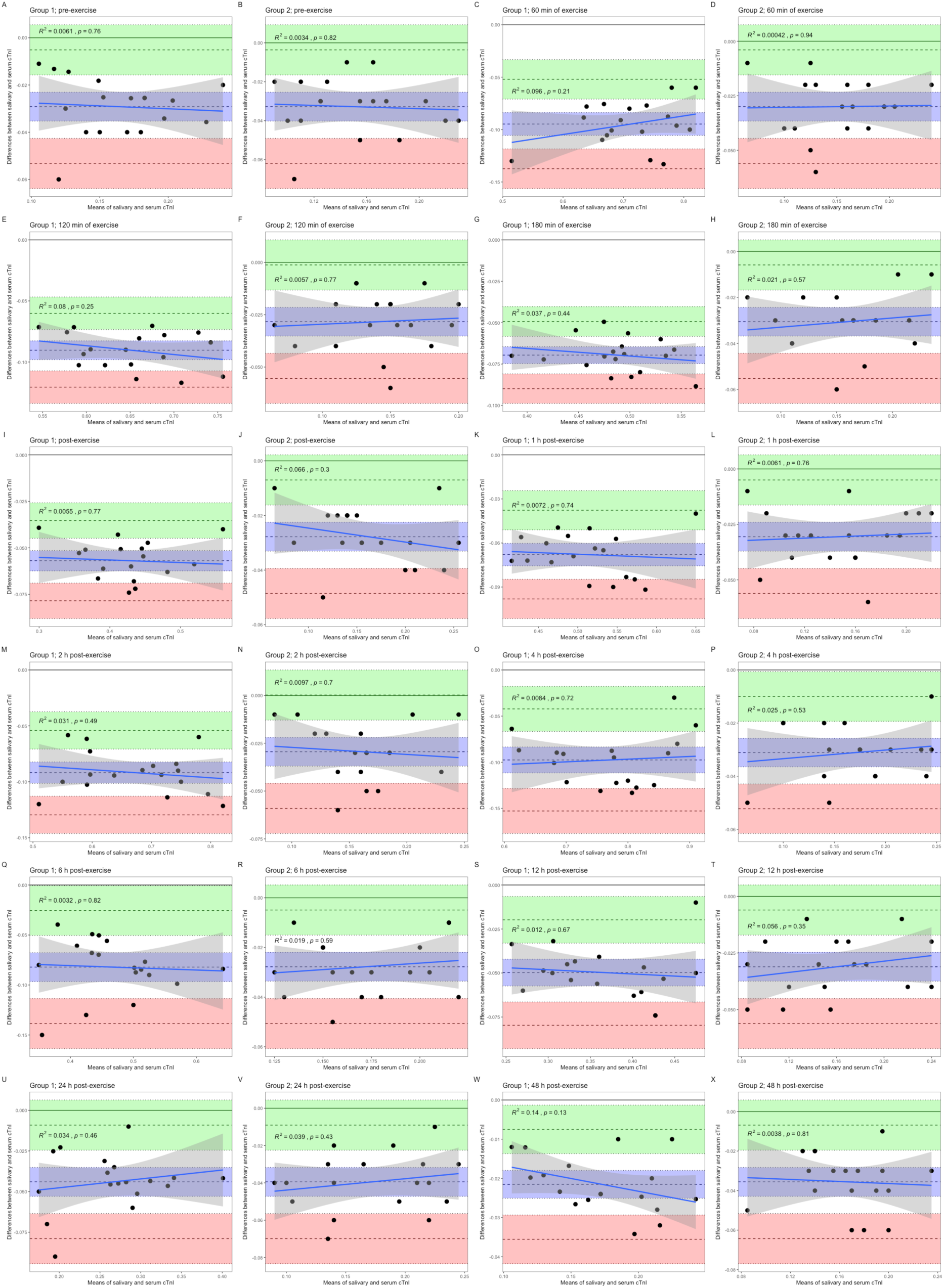
Bland–Altman plots of differences against means for comparisons of salivary and serum levels of cTnI in participants of group 1. (A, C, E, G, I, K, M, O, Q, S, U, W) and group 2 (B, D, F, H, J, L, N, P, R, T, V, X) during and after marathon running. The mean difference is represented by a dashed line (inside the blue area) parallel to the *x* axis. The limits of agreement are represented by dashed lines parallel to the *x* axis at −1.96 SD (inside the red area) and +1.96 SD (inside the green area). Shaded areas represent the 95% confidence interval limits for the mean difference (blue shading) and for the agreement limits (red and green shading). The solid blue lines represent the ordinary least squares regression of differences on means. The grey shaded regions indicate 95% confidence intervals. cTnI, cardiac troponin-I.

However, there was no proportional bias in the Bland–Altman plots, since the slope of the OLS regression of differences on means did not differ significantly from 0 (p > 0.05) and, equivalently, the 95% CI for the slope contained 0. Therefore, the limits of agreement were constructed without first finding a suitable transformation of the data. All data points were observed within ±1.96 SD of the mean difference.

## Discussion

This study is the first to examine the kinetics of cTnI release during and after marathon running using repeated blood draws and simultaneous saliva collections, aiming to compare exercise-induced changes in serum and salivary cTnI concentrations. In the present study, we revealed a biphasic response in cTnI elevations in both serum and saliva during and after marathon running. During the marathon, at approximately 60 minutes of race time, serum and salivary cTnI levels first peaked in all participants in group 1. Between 120 minutes of marathon running and the end of the race, cTnI concentrations consistently decreased in both serum and saliva. Before cTnI levels returned to baseline by 48 hours after marathon completion, a second, higher peak in serum and salivary cTnI concentrations was observed 4 hours post-exercise in all participants in group 1. Our results correspond to the findings of the previous study that reported a pattern of biphasic cTnT release into the circulation during and after a marathon run [14]. The underlying mechanisms of such biphasic release warrant consideration. Exercise-induced increase in cardiomyocyte sarcolemmal permeability seems to be primarily responsible for cTnI release within the first 60 minutes of marathon running [3, 14, 17]. Intact cTnI may penetrate from the cytosolic compartment of cardiomyocytes, particularly the free unbound pool, to the extracellular space and subsequently into the circulation through local membrane damage, exocytosis, vesicle release, and/or passive diffusion [3, 18, 19]. Mechanical stress on the cardiomyocyte and oxidative damage to its plasma membrane lipids via increased production of reactive oxygen species typically accompanying prolonged strenuous exercise might compromise membrane integrity, leading to cell wounds, increased exocytosis rates, and/or release of extracellular blebs [1, 3, 20]. In addition, when cardiomyocytes are stretched during repetitive cardiac cycles, the connection between the integrins and cytoskeleton may be disrupted, leading to the release of cTnI molecules from viable cardiomyocytes [3, 20]. Moreover, exercise-induced metabolic disturbances, including intracellular calcium overload, altered acid-base balance, and excessive waste product accumulation, might lead to reversible myocardial damage, allowing cytosolic cTnI to “leak” out [3, 20]. Therefore, significant cTnI release within the first 60 minutes of marathon running suggests that cTnI elevations are not necessarily limited to prolonged endurance exercise, but rather begin early in response to increased myocardial demand [13, 14].

A second, higher peak observed at 4 hours post-exercise seems to mainly reflect the release of fragmented cTnI rather than intact molecules [3]. Calpain-mediated cTnI proteolysis and cTnI release independently of ischemia were demonstrated on isolated rat hearts exposed to increased preload, suggesting that an acute rise in left ventricular preload during the marathon might trigger stretch-induced cardiomyocyte apoptosis [1, 21]. Furthermore, acute hemodynamic overload may also lead to cardiomyocyte apoptosis in the absence of ischemia, contributing to cTnI elevations [22]. On the other hand, there is direct evidence that brief ischemia, a state that may occur during exercise, leads to a delayed onset of cTnI release, with significant cTnI elevations beginning 30 minutes after reperfusion and persisting up to 24 hours [23]. Concomitant temporary increase in apoptosis rate was observed, with a 6-fold rise 1 hour after reperfusion followed by a 24-hour normalization [23]. Another study revealed that transient ischemia causes elevations in cTnI concentrations, which continue to consistently increase for 4 hours after ischemia [24]. Simultaneously, the magnitude and time course of cTnI elevations were dependent on ischemia duration [24]. Ischemia is known to produce degradation of cTn complexes, making fragmented cTnI more readily able to pass through the membrane [1]. For instance, ischemia provoked cTnI fragmentation in isolated rat hearts, reducing molecular mass from 24 to 15 kDa [25]. The newly available, highly sensitive troponin assays detected smaller cTnI fragments (15-20 kDa) in the circulation after exercise, distinct from the larger binary complexes or intact cTnI molecules typically identified in patients with acute myocardial infarction (AMI) [1, 19]. Therefore, brief, silent episodes of ischemia during exercise might potentially induce isolated apoptosis with the subsequent release of fragmented cTnI from both cytosolic and structurally bound pools of the cardiomyocyte into the circulation. Although apoptosis should not contribute to post-exercise cTnI elevations outside the cardiomyocyte because intracellular content is not released once the apoptotic cell is broken down and absorbed by other cells, cTnI might be released via destruction of apoptotic bodies or transition from cardiomyocyte apoptosis to secondary necrosis [26]. Additionally, it should be noted that exercise accelerates cardiomyocyte turnover [1]. The findings showed that exercise triggers the human heart’s endogenous regenerating capacity, proposing that replaced cardiomyocytes might release cTnI into the circulation if an acute bout of exercise stimulates this process [27].

Even though most exercise-induced cTnI elevations in the circulation are probably attributable to transient alterations in cardiomyocyte sarcolemmal permeability, irreversible damage associated with myocardial necrosis cannot be entirely ruled out [1]. Cardiac magnetic resonance imaging studies of participants in the Manitoba, Detroit, and London marathons revealed cTnI elevations without accompanying myocardial edema or late gadolinium enhancement [28, 29]. However, exercise-induced myocardial injury might occur at a microscopic level that is often invisible by conventional cardiac imaging techniques due to insufficient sensitivity [1]. Recent research demonstrated that cTnI concentrations exceed the upper reference limit in response to just 40 mg of myocardial necrosis in rats, but this small degree of necrosis would fail to be detected by cardiac magnetic resonance imaging [30]. Furthermore, long-term exercise training may result in cumulative cardiac micro-injury from repeated single exercise sessions [1]. This theory is supported by the fact that life-long endurance athletes have more late gadolinium enhancement compared to physically inactive individuals, and that the amount increases with years of training [31]. Certain research identified tiny patches of myocardial fibrosis in veteran athletes competing for a longer duration, located near the septum and right ventricular insertion points – areas subjected to the greatest mechanical stress during exercise [32]. Thus, although there is no concrete evidence of myocardial necrosis following exercise, it cannot be excluded as a possible cause, contributing to post-exercise cTnI elevations in some cases [1]. In the present study, the rapid time course of cTnI elevations, relatively low absolute concentrations, and return to baseline within 48 hours after marathon completion argue against irreversible damage attributable to cardiomyocyte necrosis, at least as the primary mechanism for cTnI release.

A number of alternative factors might influence cTnI levels in both serum and saliva during and after the marathon. First, exercise-induced dehydration might affect serum and salivary concentrations of cTnI, but the percentage change in fluid balance markers is typically (very) minor in comparison to the magnitude of cTnI elevations, especially post-exercise [1, 20]. It is suggested that exercise-induced dehydration is higher towards the completion of a marathon, which is contradictory to the observed decreases in cTnI levels in both serum and saliva between the first 60 minutes of marathon running and the end of the race. In addition, post-exercise rehydration is expected to quickly restore any dehydration caused by the marathon, which is also inconsistent with the progressive increases in serum and salivary cTnI concentrations up to 4 hours after exercise. Furthermore, given that hemodilution might occur after marathon completion, hemoconcentration’s effect on post-exercise cTnI levels in blood serum is probably minimal if not negligible [33]. Second, prolonged strenuous exercise and dehydration are associated with attenuated kidney function and temporarily reduced glomerular filtration rate [1, 20]. However, an increase in cTnI concentrations after a marathon run usually far exceeds the mild attenuation in renal function, suggesting that the contribution of reduced renal function to the magnitude of post-exercise cTnI elevations in the circulation is non-significant [1, 34]. Third, it was proposed that either skeletal muscle damage with cTnI release or the assay’s cross-reactivity with skeletal Tn might lead to overestimation of cTnI levels [1]. Available data contradict this hypothesis because cTnT but not cTnI was identified in the samples of healthy human skeletal muscle [35]. Similarly, in patients with skeletal myopathies, serum cTnT concentrations often exceeded the upper reference limit, whereas serum cTnI levels were rarely elevated [36]. Moreover, we are not aware of any findings demonstrating cTnI elevations in skeletal muscle samples from exercising individuals. Therefore, skeletal muscle damage caused by exercise might conceivably contribute to cTnT elevations in the circulation but is unlikely to contribute to cTnI elevations [1].

Importantly, our data are the first to demonstrate that there was a similar pattern of time-dependent alterations in cTnI levels in serum and saliva during and after marathon running, as evidenced by cTnI quantifications across twelve time points. Although there was a proportional agreement between serum and salivary cTnI concentrations in both groups at all twelve time points, there was also a negative differential bias of the new measurement method (i.e., measurement of salivary cTnI concentrations) compared to the reference standard (i.e., measurement of serum cTnI concentrations). In other words, cTnI concentrations in blood serum were consistently higher than those in saliva in both groups at all twelve time points, corroborating a previous report [13]. A strong correlation between serum and salivary levels of cTnI aligns with and extends recent work, confirming that exercise-induced changes in salivary cTnI concentrations may faithfully reflect serum cTnI kinetics, even across a significantly longer and more metabolically complex exercise stimulus [13]. Mechanisms underlying the transfer of cTnI across the blood-salivary barrier remain unclear. The ability of intact cTnI molecules to cross the glomerular filtration barrier was demonstrated by recent clinical trials employing a highly sensitive cTnI assay [37, 38]. This implies that a parallel transport mechanism might mediate the transfer of cTnI into saliva across the blood-salivary barrier [13]. Passive diffusion of cTnI from the bloodstream to saliva could represent one route by which intact cTnI from an early releasable pool and cTnI fragments are cleared from the circulation [37]. This hypothesis is supported by the fact that a (very) low background level of cTnI was detected in the saliva of the healthy population using a highly sensitive cTnI assay capable of identifying single cTnI molecules [39]. It is possible, therefore, that there is a continual turnover of cTnI and that exercise is a stimulus for an accelerated turnover, facilitating the transport of cTnI across the blood-salivary barrier due to increases in cell membrane permeability [13]. Such increases in permeability are most likely caused by elevated production of reactive oxygen species, leading to oxidative damage to plasma membrane lipids in vascular endothelial and/or salivary gland cells [40]. All things considered, it is plausible that reversible cell membrane damage might be mainly responsible for cTnI elevations in saliva during and after the marathon.

One possible limitation that should be noted here is that the findings are difficult to extrapolate to all athletes involved in different sports because only well-trained male runners participated in this study.

## Conclusions and perspectives

Finally, we have developed and validated a non-invasive approach for mapping the detailed kinetics of cTnI release induced by prolonged strenuous exercise. Documenting a similar, biphasic pattern of cTnI elevations in saliva and serum during and after the marathon provides a reliable non-invasive alternative without requiring a blood draw, which may aid in assessing whether exercise-induced cTnI elevations are transient or sustained. For the sports and exercise medicine setting, the portability and non-invasiveness of POC cTnI testing in saliva enable potential applications in field-based monitoring of cTnI elevations during training sessions and/or competition races, and tracking of individual post-exercise cTnI kinetics outside the laboratory to inform return-to-exercise decisions. Additionally, the suggested saliva-based cTnI assay creates new opportunities for the development of a biosensor for ongoing monitoring of the magnitude and time course of cTnI elevations caused by strenuous exercise in vulnerable individuals. Large prospective studies with extended follow-up in clinical and recreational athletic populations are needed to determine whether exercise-induced cTnI elevations in saliva may predict future cardiovascular events. Subsequent studies can then determine if a specific cut-off value (i.e., the 99th percentile of the upper reference limit) for salivary cTnI may differentiate physiological and reversible from pathological and irreversible myocardial injury, becoming an important adjunct to AMI diagnostic protocols.

## Declarations

### Ethics approval and consent to participate

This study was approved by the Local Ethics Committee of Lobachevsky University (Nr. 2, 22.09.2025) and was conducted in accordance with the principles set forth in the Declaration of Helsinki. Informed consent was obtained from all individuals included in this study.

## Consent for publication

All participants consented to publication of the findings.

## Availability of data and material

The data that support the findings of this study are available on request from the corresponding author.

## Authors’ contributions

**AO:** Conceptualization; Data curation; Formal analysis; Funding acquisition; Investigation; Methodology; Resources; Visualization; Writing – original draft; Writing – review & editing. **AP:** Writing – original draft; Writing – review & editing.

The authors have accepted responsibility for the entire content of this manuscript and approved its submission.

## Abbreviations

AMI: Acute myocardial infarction
CI: Confidence interval
cTn: Cardiac troponin
cTnI: Cardiac troponin-I
cTnT: Cardiac troponin-T
OLS: Ordinary least squares
POC: Point-of-care
SD: Standard deviation

## Acknowledgments

The authors appreciate male runners and their coaches for their collaboration and commitment to this study.

## References

1. Aengevaeren VL, Baggish AL, Chung EH, et al. Exercise-Induced Cardiac Troponin Elevations: From Underlying Mechanisms to Clinical Relevance. Circulation. 2021;144(24):1955–72.

2. Omland T, Aakre KM. Cardiac Troponin Increase After Endurance Exercise. Circulation. 2019;140(10):815–8.

3. Shave R, Baggish A, George K, Wood M, Scharhag J, Whyte G, Gaze D, Thompson PD. Exercise-induced cardiac troponin elevation: evidence, mechanisms, and implications. J Am Coll Cardiol. 2010;56(3):169–76.

4. Jean-Gilles M, Baggish A. Cardiac troponin elevation in athletes: blame the musician and not the instrument. Br J Sports Med. 2024;58:121–2.

5. Aengevaeren VL, Hopman MTE, Thompson PD, et al. Exercise-Induced Cardiac Troponin I Increase and Incident Mortality and Cardiovascular Events. Circulation. 2019;140(10):804–14.

6. Pelliccia A, Solberg EE, Papadakis M, et al. Recommendations for participation in competitive and leisure time sport in athletes with cardiomyopathies, myocarditis, and pericarditis: position statement of the Sport Cardiology Section of the European Association of Preventive Cardiology (EAPC). Eur Heart J. 2019;40(1):19–33.

7. Anzini M, Merlo M, Sabbadini G, et al. Long-term evolution and prognostic stratification of biopsy-proven active myocarditis. Circulation. 2013;128(22):2384–94.

8. Boeddinghaus J, Nestelberger T, Koechlin L, et al. Early Diagnosis of Myocardial Infarction with Point-of-Care High-Sensitivity Cardiac Troponin I. J Am Coll Cardiol. 2020;75(10):1111–24.

9. Titus J, Wu AHB, Biswal S, Burman A, Sengupta SP, Sengupta PP. Development and preliminary validation of infrared spectroscopic device for transdermal assessment of elevated cardiac troponin. Commun Med (Lond). 2022;2:42.

10. Lindsay A, Costello JT. Realising the Potential of Urine and Saliva as Diagnostic Tools in Sport and Exercise Medicine. Sports Med. 2017;47(1):11–31.

11. Floriano PN, Christodoulides N, Miller CS, et al. Use of saliva-based nano-biochip tests for acute myocardial infarction at the point of care: a feasibility study. Clin Chem. 2009;55(8):1530–8.

12. Herr AE, Hatch AV, Throckmorton DJ, Tran HM, Brennan JS, Giannobile WV, Singh AK. Microfluidic immunoassays as rapid saliva-based clinical diagnostics. Proc Natl Acad Sci. 2007;104(13):5268–73.

13. Ovchinnikov AN. Utilizing saliva for non-invasive detection of exercise-induced myocardial injury with point-of-care cardiac troponin-I. Sci Rep. 2025;15(1):27283.

14. Middleton N, George K, Whyte G, Gaze D, Collinson P, Shave R. Cardiac troponin T release is stimulated by endurance exercise in healthy humans. J Am Coll Cardiol. 2008;52(22):1813–4.

15. World Medical Association Declaration of Helsinki. Recommendations guiding physicians in biomedical research involving human subjects. JAMA. 1997;277:925–6.

16. Burke FJ, Busby M, McHugh S, Delargy S, Mullins A, Matthews R. Evaluation of an oral health scoring system by dentists in general dental practice. Br Dent J. 2003;194(4):215–8.

17. Aengevaeren VL, Froeling M, Hooijmans MT, et al. Myocardial Injury and Compromised Cardiomyocyte Integrity Following a Marathon Run. JACC Cardiovasc Imaging. 2020;13(6):1445–7.

18. Airaksinen KEJ. Cardiac Troponin Release After Endurance Exercise: Still Much to Learn. J Am Heart Assoc. 2020;9(4):e015912.

19. Celeski M, Segreti A, Crisci F, et al. The Role of Cardiac Troponin and Other Emerging Biomarkers Among Athletes and Beyond: Underlying Mechanisms, Differential Diagnosis, and Guide for Interpretation. Biomolecules. 2024;14(12):1630.

20. Gresslien T, Agewall S. Troponin and exercise. Int J Cardiol. 2016;221:609–21.

21. Feng J, Schaus BJ, Fallavollita JA, Lee TC, Canty JM Jr. Preload induces troponin I degradation independently of myocardial ischemia. Circulation. 2001;103(16):2035–7.

22. Weil BR, Suzuki G, Young RF, Iyer V, Canty JM Jr. Troponin Release and Reversible Left Ventricular Dysfunction After Transient Pressure Overload. J Am Coll Cardiol. 2018;71(25):2906–16.

23. Weil BR, Young RF, Shen X, Suzuki G, Qu J, Malhotra S, Canty JM Jr. Brief Myocardial Ischemia Produces Cardiac Troponin I Release and Focal Myocyte Apoptosis in the Absence of Pathological Infarction in Swine. JACC Basic Transl Sci. 2017;2(2):105–14.

24. Árnadóttir Á, Pedersen S, Bo Hasselbalch R, Goetze JP, Friis-Hansen LJ, Bloch-Münster AM, Skov Jensen J, Bundgaard H, Iversen K. Temporal Release of High-Sensitivity Cardiac Troponin T and I and Copeptin After Brief Induced Coronary Artery Balloon Occlusion in Humans. Circulation. 2021;143(11):1095–104.

25. McDonough JL, Arrell DK, Van Eyk JE. Troponin I degradation and covalent complex formation accompanies myocardial ischemia/reperfusion injury. Circ Res. 1999;84(1):9–20.

26. Hammarsten O, Mair J, Möckel M, Lindahl B, Jaffe AS. Possible mechanisms behind cardiac troponin elevations. Biomarkers. 2018;23(8):725–34.

27. Vujic A, Lerchenmüller C, Wu TD, et al. Exercise induces new cardiomyocyte generation in the adult mammalian heart. Nat Commun. 2018;9(1):1659.

28. Mousavi N, Czarnecki A, Kumar K, et al. Relation of biomarkers and cardiac magnetic resonance imaging after marathon running. Am J Cardiol. 2009;103(10):1467–72.

29. O’Hanlon R, Wilson M, Wage R, et al. Troponin release following endurance exercise: is inflammation the cause? a cardiovascular magnetic resonance study. J Cardiovasc Magn Reson. 2010;12(1):38.

30. Marjot J, Kaier TE, Martin ED, et al. Quantifying the Release of Biomarkers of Myocardial Necrosis from Cardiac Myocytes and Intact Myocardium. Clin Chem. 2017;63(5):990–6.

31. van de Schoor FR, Aengevaeren VL, Hopman MT, Oxborough DL, George KP, Thompson PD, Eijsvogels TM. Myocardial Fibrosis in Athletes. Mayo Clin Proc. 2016;91(11):1617–31.

32. La Gerche A. Can intense endurance exercise cause myocardial damage and fibrosis? Curr Sports Med Rep. 2013;12(2):63–9.

33. Neumayr G, Pfister R, Hoertnagl H, Mitterbauer G, Prokop W, Joannidis M. Renal function and plasma volume following ultramarathon cycling. Int J Sports Med. 2005;26(1):2–8.

34. Wołyniec W, Ratkowski W, Renke J, Renke M. Changes in Novel AKI Biomarkers after Exercise. A Systematic Review. Int J Mol Sci. 2020;21(16):5673.

35. Ricchiuti V, Apple FS. RNA expression of cardiac troponin T isoforms in diseased human skeletal muscle. Clin Chem. 1999;45(12):2129–35.

36. Schmid J, Liesinger L, Birner-Gruenberger R, et al. Elevated Cardiac Troponin T in Patients with Skeletal Myopathies. J Am Coll Cardiol. 2018;71(14):1540–9.

37. Chaulin AM. Metabolic Pathway of Cardiospecific Troponins: From Fundamental Aspects to Diagnostic Role (Comprehensive Review). Front Mol Biosci. 2022;9:841277.

38. Chen JY, Lee SY, Li YH, Lin CY, Shieh MD, Ciou DS. Urine High-Sensitivity Troponin I Predict Incident Cardiovascular Events in Patients with Diabetes Mellitus. J Clin Med. 2020;9(12):3917.

39. Chekin F, Vasilescu A, Jijie R, et al. Sensitive electrochemical detection of cardiac troponin I in serum and saliva by nitrogen-doped porous reduced graphene oxide electrode. Sensors Actuators B Chem. 2018;262:18–7.

40. Ovchinnikov AN, Paoli A, Seleznev VV, Deryugina AV. Measurement of Lipid Peroxidation Products and Creatine Kinase in Blood Plasma and Saliva of Athletes at Rest and following Exercise. J Clin Med. 2022;11(11):3098.

